## Supplementary material for "Transcriptional and spatial profiling of the kidney allograft unravels a central role for FcyRIII+ innate immune cells in rejection": Supplementary Information 07.07.22.pdf

**Supplemental Material and Methods**

**Multiplex immunofluorescence staining and image processing (Opal)**

A multiplex immunofluorescence staining method on paraffin-embedded tissue was recently described using Opal reagents (PerkinElmer, Waltham, MA).[1] Briefly, tissue sections were deparaffinized, rehydrated and fixed for 20 minutes in 10% neutral-buffered formalin. Antigen retrieval was performed using microwave treatment (MWT) in antigen retrieval solution pH6 or pH9 (AR6 or AR9) according to the target of interest. At each of 4 consecutive staining cycles, primary antibodies anti-NKp46 (clone 8E5B,17 Innate Pharma), anti-CD163 (clone 10D6, Leica Biosystems), anti-CD34 (clone QBEnd-10, Dako), and anti-CD3 (clone SP7, Thermo scientific) were added, followed by Opal Polymer HRP Ms + Rb Kit (PerkinElmer) for 10 minutes at room temperature. The tissue sections were then incubated with TSA opal fluorophores (Opal 620, 690, 520 and 540). After each staining cycle, MWT was performed to remove antibody-TSA complex with AR6 or AR9. Finally, all slides were counterstained with DAPI for 5 minutes.

The tissue slides were initially scanned using the PerkinElmer Vectra (v3.0; PerkinElmer) at low magnification (×10). Under pathologist supervision, regions of interest (ROI) were identified and scanned at high resolution (x20) using the Phenochart 1.0.4 viewer (PerkinElmer). High resolution scans of ROIs in the 86 biopsies were analyzed using the software inForm Tissue Finder 2.3.0 (PerkinElmer). The consecutive steps in the image processing software were tissue segmentation based on CD34 (discerning the extravascular from the intravascular compartment), nuclear segmentation based on DAPI staining and cell phenotyping based on CD163, NKp46 and CD3, resulting in a label as “Macrophage”, “NK cell”, “T cell” and “Other” for each of the identified cells. Infiltration of the respective immune cell types was quantified as density, i.e. number of cell per mm², and relative prevalence, i.e. the number of a specific immune cell type compared to the total number of Macrophages, NK cells and T cells.

**Supplementary tables**

Table S1. Clinical and pathological features of the scRNAseq study population. DBD: donation after brain death, DCD: donation after circulatory death, LD: living donation, M: male, F: female, HLA-DSA: donor-specific anti-human leukocyte antigen antibodies, NR: no rejection, ABMR: antibody-mediated rejection, TCMR: T cell-mediated rejection

| **Biopsy no.** | **Recipient age range at tx** | **Indication/ protocol** | **Days after tx** | **Donation type** | **Recipient and donor gender** | **Induction** | **Serum creatinine (mg/dl)** | **HLA-DSA** | **Histological lesions** | **Biopsy group according to Vaulet et al[2]** |
|---|---|---|---|---|---|---|---|---|---|---|
| #1 | 31-35 | Indication | 37 | DBD | M/M | Basiliximab | 1.76 | A11 | g0, ptc0, C4d0, v0, cg0, t0, i0, ct0, ci0 | NR DSA+ |
| #2 | 51-55 | Indication | 61 | DCD | F/F | None | 4.46 | DPB1*03:01 (104:01) | g2, ptc2, C4d2, v0, cg0, t0, i1, ct1, ci0 | ABMR |
| #3 | 41-45 | Indication | 12 | LD | M/F | Basiliximab | 2.09 | DQ1 (DQA1*01:04/ DQB1*05) | g0, ptc0, C4d0, v0, cg0, t1, i1, ct0, ci0 | NR DSA+ |
| #4 | 51-55 | Protocol | 91 | DCD | F/F | None | 1.92 | DPB1*03:01 (104:01) | g1, ptc1, C4d1, v0, cg0, t0, i0, ct0, ci0 | NR DSA+ |
| #5 | 56-60 | Indication | 9 | DBD | M/F | Basiliximab | 1.86 | B62, B35 | g1, ptc2, C4d0, v1, cg0, t0, i0, ct0, ci0 | NR DSA+ |
| #6 | 11-15 | Indication | 7709 | DBD | M/M | Thymoglobulin | 8.08 | No HLA-DSA | g0, ptc0, C4d0, v0, cg0, t0, i0, ct2, ci2 | NR DSA- |
| #7 | 51-55 | Indication | 6 | DBD | M/M | Basiliximab | 6.93 | DRB1*08:02 | g2, ptc2, C4d1, v1, cg0, t1, i1, ct1, ci1 | ABMR |
| #8 | 61-65 | Indication | 6 | DCD | F/F | None | 5.03 | No HLA-DSA | g0, ptc2, C4d1, v0, cg0, t1, i1, ct0, ci0 | TCMR |
| #9 | 16-20 | Protocol | 91 | LD | M/M | Basiliximab | 1.04 | No HLA-DSA | g0, ptc0, C4d1, v0, cg0, t0, i0, ct0, ci0 | NR DSA- |
| #10 | 56-60 | Indication | 44 | DCD | M/F | Basiliximab | 3.94 | No HLA-DSA | g0, ptc0, C4d1, v0, cg0, t0, i0, ct1, ci1 | NR DSA- |
| #11 | 51-55 | Indication | 247 | DCD | F/F | Basiliximab | 1.87 | DQ1 (DQB1*05:01 DQA1*01:01) | g0, ptc0, C4d0, v0, cg0, t0, i0, ct1, ci1 | NR DSA+ |

| | | | | | | | | | | |
|---|---|---|---|---|---|---|---|---|---|---|
| #12 | 46-50 | Indication | 2104 | DCD | M/M | Basiliximab | 2.64 | A32 | g1, ptc0, C4d0, v0, cg0, t0, i0, ct1, ci1 | NR DSA+ |
| #13 | 61-65 | Indication | 118 | DCD | M/F | Basiliximab | 2.27 | A23, DR7 | g0, ptc0, C4d2, v0, cg0, t1, i1, ct1, ci1 | NR DSA+ |
| #14 | 46-50 | Indication | 6 | DBD | M/F | Basiliximab | 6.23 | No HLA-DSA | g0, ptc0, C4d0, v0, cg0, t1, i1, ct1, ci1 | NR DSA- |
| #15 | 51-55 | Indication | 361 | DCD | F/F | Basiliximab | 3.10 | DQ1 (DQB1*05:01 DQA1*01:01) | g0, ptc1, C4d0, v0, cg0, t1, i1, ct2, ci2 | NR DSA+ |
| #16 | 21-25 | Indication | 2002 | DBD | F/M | Basiliximab | 1.93 | Cw6 DQ5 | g2, ptc2, C4d3, v0, cg1, t0, i0, ct2, ci2 | ABMR |

Table S2. Clinical and pathological features of the Multiple Iterative Labeling by Antibody Neodeposition (MILAN) study population

| **Biopsy no.** | **Recipient age range at tx** | **Indication /protocol** | **Days** after **tx** | **Donation type** | **Recipient and donor gender** | **Induction** | **Serum creatinine (mg/dl)** | **HLA-DSA** | **Histological lesions** | **Biopsy group according to Vaulet et al**[2] |
|---|---|---|---|---|---|---|---|---|---|---|
| #1 | 66-70 | Indication | 16 | DBD | M/F | Basiliximab | 4.47 | No HLA-DSA | g3, ptc1, C4d1, v0, cg0, t1, i0, ct0, ci0 | ABMR DSA- |
| #2 | 36-40 | Indication | 5 | DBD | M/M | Basiliximab | 6.92 | No HLA-DSA | g3, ptc2, C4d3, v0, cg0, t3, i2, ct0, ci0 | ABMR DSA- |
| #3 | 56-60 | Indication | 70 | LD | M/F | Basiliximab | 2.87 | DR7 | g0, ptc0, C4d0, v0, cg0, t0, i0, ct1, ci0 | NR DSA+ |
| #4 | 36-40 | Indication | 19 | DBD | M/M | Basiliximab | 2.33 | No HLA-DSA | g3, ptc1, C4d3, v0, cg0, t1, i0, ct0, ci0 | ABMR DSA- |
| #5 | 26-30 | Protocol | 96 | DBD | F/F | Basiliximab | 0.92 | B60<br>DQA1 | g3, ptc0, C4d3, v0, cg0, t0, i0, ct1, ci0 | ABMR |
| #6 | 56-60 | Indication | 59 | DBD | M/M | None | 3.65 | No HLA-DSA | g0, ptc0, C4d0, v0, cg0, t0, i0, ct1, ci0 | NR DSA- |
| #7 | 21-25 | Indication | 276 | DBD | M/M | Basiliximab | 1.45 | DPB1*03:01<br>DPB1*04:01 | g2, ptc3, C4d0, v0, cg1, t1, i2, ct1, ci0 | ABMR |
| #8 | 71-75 | Indication | 6 | DBD | M/F | None | 3.95 | No HLA-DSA | g0, ptc0, C4d0, v1, cg0, t2, i2, ct1, ci0 | TCMR |
| #9 | 41-45 | Indication | 6 | DBD | M/F | Basiliximab | 10.23 | A2 | g2, ptc2, C4d1, v1, cg0, t2, i3, ct1, ci1 | Mixed |
| #10 | 26-30 | Protocol | 1825 | DBD | M/M | Basiliximab | 1.46 | No HLA-DSA | g0, ptc0, C4d0, v0, cg0, t2, i2, ct1, ci1 | TCMR |
| #11 | 41-45 | Protocol | 371 | LD | F/F | Basiliximab | 1.70 | No HLA-DSA | g0, ptc0, C4d0, v0, cg0, t2, i2, ct0, ci0 | TCMR |
| #12 | 51-55 | Protocol | 363 | LD | F/M | Basiliximab | 2.15 | No HLA-DSA | g2, ptc1, C4d0, v1, cg0, t1, i1, ct1, ci0 | ABMR DSA- |
| #13 | 56-60 | Protocol | 374 | DBD | F/F | Basiliximab | 1.13 | No HLA-DSA | g2, ptc2, C4d0, v0, cg0, t0, i0, ct2, ci2 | ABMR DSA- |

| | | | | | | | | | | |
|---|---|---|---|---|---|---|---|---|---|---|
| #14 | 46-50 | Indication | 9 | DBD | F/F | Basiliximab | 3.08 | No HLA-DSA | g3, ptc0, C4d1, v1, cg0, t2, i3, ct0, ci0 | ABMR DSA- |
| #15 | 56-60 | Indication | 172 | DCD | F/M | Basiliximab | 2.41 | DPA1*02 | g3, ptc0, C4d0, v1, cg0, t2, i3, ct0, ci0 | Mixed |
| #16 | 66-70 | Indication | 5 | DBD | M/F | None | 1.80 | No HLA-DSA | g0, ptc0, C4d0, v0, cg0, t2, i3, ct0, ci0 | TCMR |
| #17 | 51-55 | Indication | 61 | DCD | F/F | None | 4.46 | DPB1*03:01 (104:01) | g2, ptc2, C4d2, v0, cg0, t0, i1, ct1, ci0 | ABMR |
| #18 | 16-20 | Protocol | 91 | LD | M/M | Basiliximab | 1.04 | No HLA-DSA | g0, ptc0, C4d1, v0, cg0, t0, i0, ct0, ci0 | NR DSA- |

Table S3. Antibodies used for MILAN. AF; Alexa Fluor

| Marker | Analysis | Reference | Supplier | Host species | Clone | Dilution |
|---|---|---|---|---|---|---|
| AQP1 | Phenotypic identification | AB2219 | Sigma Aldrich | Rabbit | Polyclonal | 1:20000 |
| CD1c | Phenotypic identification/Monocyte subclustering | UM500042 | OriGene | Mouse IgG1 | UMAB46 | 1:333 |
| CD3 | Phenotypic identification | MA1-90582 | ThermoFisher | Rabbit | SP7 | 1:200 |
| CD4 | Phenotypic identification | ab133616 | Abcam | Rabbit | EPR6855 | 1:166 |
| CD8 | Phenotypic identification | sc-53212 | Santa Cruz Biotechnology | Mouse IgG1 | C8/144B | 1:500 |
| CD11b | Monocyte subclustering | ab133357 | Abcam | Rabbit | EPR1344 | 1:5000 |
| CD11c | Phenotypic identification/Monocyte subclustering | sc-46676 | Santa Cruz Biotechnology | Mouse IgG1 | B-6 | 1:75 |
| CD14 | Phenotypic identification/Monocyte subclustering | #75181 | Cell Signaling Technology | Rabbit | D7A2T | 1:200 |
| CD16 | Phenotypic identification/Monocyte subclustering | NCL-L-CD16 | Leica Biosystems | Mouse IgG2a | 2H7 | 1:333 |
| CD20 | Phenotypic identification | M075501-2 | Agilent | Mouse IgG2a | L26 | 1:630 |
| CD31 | Exploratory Analysis | LS-C173974 | LSBio | Mouse IgG2a | OTI2C6 | 1:1000 |
| CD56 | Phenotypic identification | sc-7326 | Santa Cruz Biotechnology | Mouse IgG1 | 123C3.D5 | 1:200 |
| CD57 | Phenotypic identification | MAB8560 | R&D Systems | Mouse IgG2a | 1002707 | 1:250 |
| CD68 | Phenotypic identification/Monocyte subclustering | MA5-12407 | ThermoFisher | Mouse IgG3 | PGM1 | 1:200 |
| CD69 | Exploratory Analysis | HPA050525 | Sigma Aldrich | Rabbit | Polyclonal | 1:50 |
| CD79a | Exploratory Analysis | sc-53209 | Santa Cruz Biotechnology | Mouse IgG1 | JCB117 | 1:2000 |
| CD123 | Monocyte subclustering | NCL-L-CD123 | Leica Biosystems | Mouse IgG2b | BR4MS | 1:10 |
| CD138 | Phenotypic identification | MCA2459GA | BioRad | Mouse IgG1 | B-A38 | 1:200 |
| CD141 | Monocyte subclustering | sc-13164 | Santa Cruz Biotechnology | Mouse IgG2a | D-3 | 1:200 |
| CD163 | Phenotypic identification/Monocyte subclustering | ab182422 | Abcam | Rabbit | EPR19518 | 1:227 |
| CD206 | Monocyte subclustering | MAB25341 | R&D Systems | Mouse IgG2b | #685645 | 1:250 |
| CD209 | Monocyte subclustering | MAB161 | R&D Systems | Mouse IgG2b | 120507 | 1:333 |
| Collagen IV | Exploratory Analysis | ab6311 | Abcam | Mouse IgG1 | COL-94 | 1:333 |
| FOXP3 | Phenotypic identification | ab20034 | Abcam | Mouse IgG1 | 236A/E7 | 1:1000 |

| | | | | | | |
|---|---|---|---|---|---|---|
| GrB | Exploratory Analysis | sc-73620 | Santa Cruz Biotechnology | Mouse IgG2a | GRB7 | 1:50 |
| HLA-DR | Phenotypic identification/Monocyte subclustering | sc-56545 | Santa Cruz Biotechnology | Mouse IgG2b | SPM289 | 1:333 |
| IRF8 | Monocyte subclustering | sc-365042 | Santa Cruz Biotechnology | Mouse IgG1 | E-9 | 1:400 |
| Ki67 | Exploratory Analysis | UM800033 | OriGene | Mouse IgG2a | UMAB107 | 1:5000 |
| LAG3 | Exploratory Analysis | #15372 | Cell Signaling Technology | Rabbit | D2G4O | 1:250 |
| MPO | Phenotypic identification | ab93665 | Abcam | Rabbit | SP72 | 1:1600 |
| OX40 | Exploratory Analysis | #61637 | Cell Signaling Technology | Rabbit | E9U7O | 1:50 |
| PanCK | Phenotypic identification | 53-9003-80 | ThermoFisher | Mouse IgG1 | AE1/AE3 | 1:1250 |
| PD-1 | Exploratory Analysis | #86163 | Cell Signaling Technology | Rabbit | D4W2J | 1:50 |
| PD-L1 | Exploratory Analysis | #13684 | Cell Signaling Technology | Rabbit | E1L3N | 1:50 |
| PRF1 | Phenotypic identification | ab89821 | Abcam | Mouse IgG1 | 5B10 | 1:20 |
| S100 | Phenotypic identification | Z0311 | Agilent | Rabbit | Polyclonal | 1:4120 |
| TCF7 | Exploratory Analysis | #2203 | Cell Signaling Technology | Rabbit | C63D9 | 1:67 |
| TIM3 | Exploratory Analysis | AF2365 | R&D Systems | Goat | Polyclonal | 1:200 |
| AF-488 anti-mouse IgG2a | Secondary antibody | 115-545-206 | Jackson Immunoresearch | Goat | Polyclonal | 5 µg/mL |
| AF-488 anti-mouse IgG2b | Secondary antibody | 115-545-207 | Jackson Immunoresearch | Goat | Polyclonal | 5 µg/mL |
| AF-488 anti-mouse IgG3 | Secondary antibody | 115-545-209 | Jackson Immunoresearch | Goat | Polyclonal | 5 µg/mL |
| AF-555 anti-mouse IgG1 | Secondary antibody | A-21127 | Invitrogen | Goat | Polyclonal | 5 µg/mL |
| AF-555 anti-mouse IgG2a | Secondary antibody | A-21137 | Invitrogen | Goat | Polyclonal | 5 µg/mL |
| AF-555 anti-mouse IgG2b | Secondary antibody | A-21147 | Invitrogen | Goat | Polyclonal | 5 µg/mL |
| AF- 555 anti-goat | Secondary antibody | A-21432 | Invitrogen | Donkey | Polyclonal | 5 µg/mL |
| AF- 647 anti-rabbit | Secondary Antibody | 711-605-152 | Jackson Immunoresearch | Donkey | Polyclonal | 5 µg/mL |

### Supplementary figures

Suppl Fig. 1

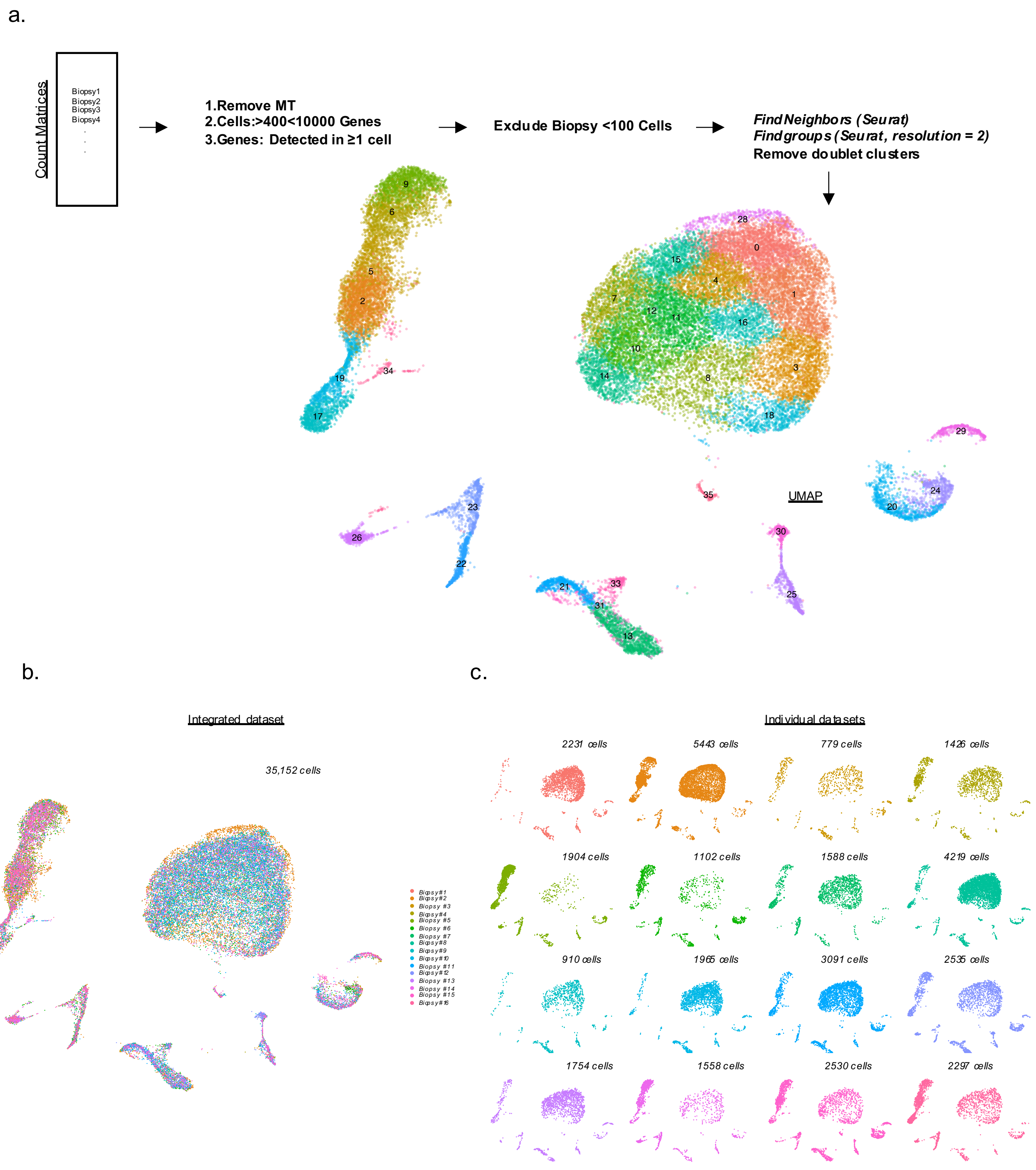
a.
Count Matrices
Biopsy1
Biopsy2
Biopsy3
Biopsy4
1.Remove MT
2.Cells:>400<10000 Genes
3.Genes: Detected in ≥1 cell
Exclude Biopsy <100 Cells
FindNeighbors (Seurat)
Findgroups (Seurat, resolution = 2)
Remove doublet clusters
UMAP
b.
Integrated dataset
35,152 cells
c.
Individual datasets
2231 cells
5443 cells
779 cells
1426 cells
1904 cells
1102 cells
1588 cells
4219 cells
910 cells
1965 cells
3091 cells
2535 cells
1754 cells
1558 cells
2530 cells
2297 cells

Suppl Fig. 2

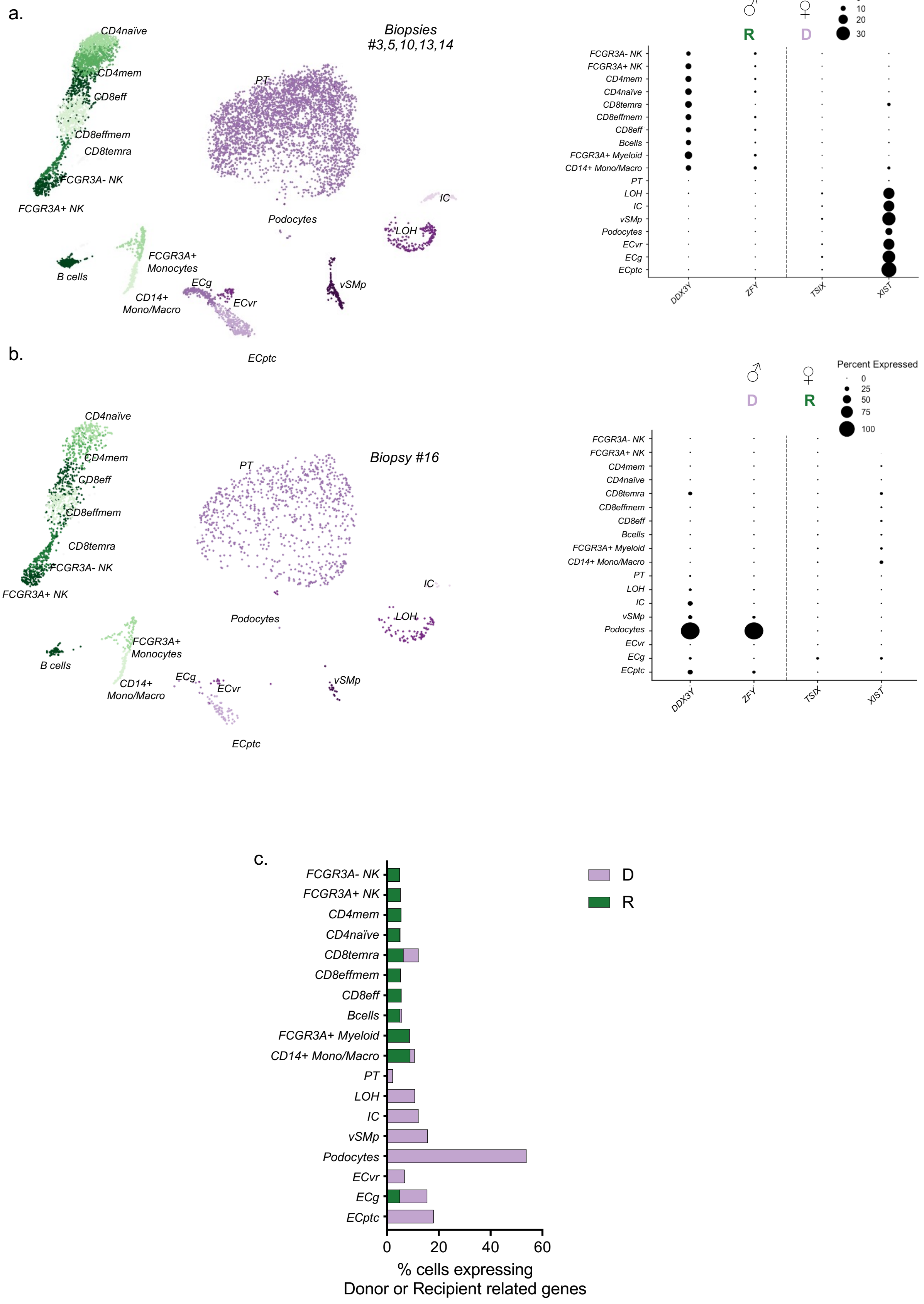
a.
Biopsies
#3,5,10,13,14
CD4naïve
CD4mem
CD8eff
CD8effmem
CD8temra
FCGR3A- NK
FCGR3A+ NK
B cells
FCGR3A+
Monocytes
CD14+
Mono/Macro
PT
Podocytes
IC
LOH
vSMp
ECg
ECvr
ECptc
R
D
Percent Expressed
0
10
20
30
FCGR3A+ Myeloid
CD14+ Mono/Macro
Bcells
DDX3Y
ZFY
TSIX
XIST
b.
Biopsy #16
D
R
Percent Expressed
0
25
50
75
100
c.
D
R
0
20
40
60
% cells expressing
Donor or Recipient related genes

Supp Fig. 3

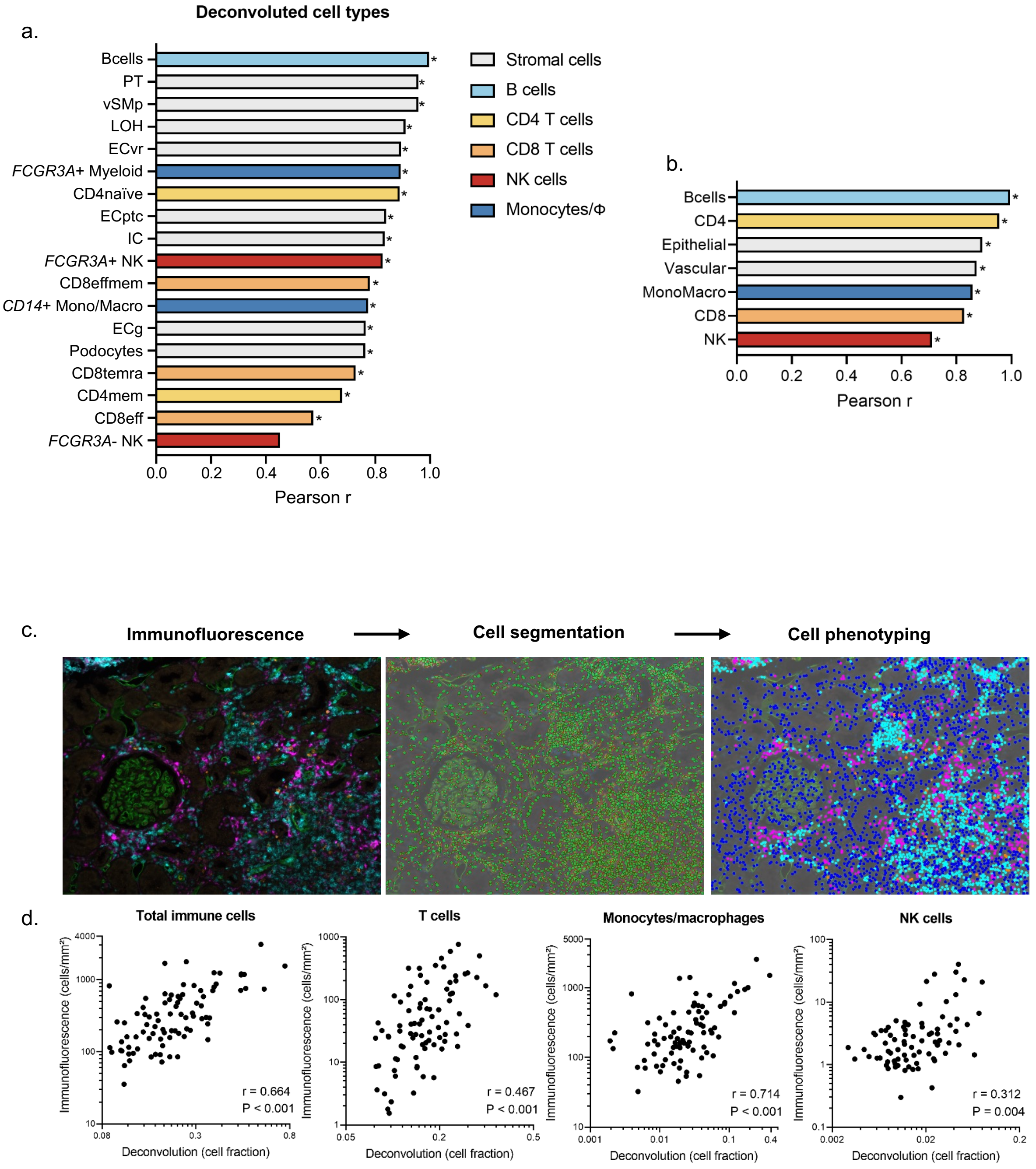

a.
Deconvoluted cell types
Bcells
PT
vSMp
LOH
ECvr
FCGR3A+ Myeloid
CD4naïve
ECptc
IC
FCGR3A+ NK
CD8effmem
CD14+ Mono/Macro
ECg
Podocytes
CD8temra
CD4mem
CD8eff
FCGR3A- NK
0.0 0.2 0.4 0.6 0.8 1.0
Pearson r
Stromal cells
B cells
CD4 T cells
CD8 T cells
NK cells
Monocytes/Φ
b.
Bcells
CD4
Epithelial
Vascular
MonoMacro
CD8
NK
0.0 0.2 0.4 0.6 0.8 1.0
Pearson r
c.
Immunofluorescence
Cell segmentation
Cell phenotyping
d.
Total immune cells
T cells
Monocytes/macrophages
NK cells
Immunofluorescence (cells/mm²)
Deconvolution (cell fraction)
r = 0.664
P < 0.001
r = 0.467
P < 0.001
r = 0.714
P < 0.001
r = 0.312
P = 0.004

Suppl Fig. 4

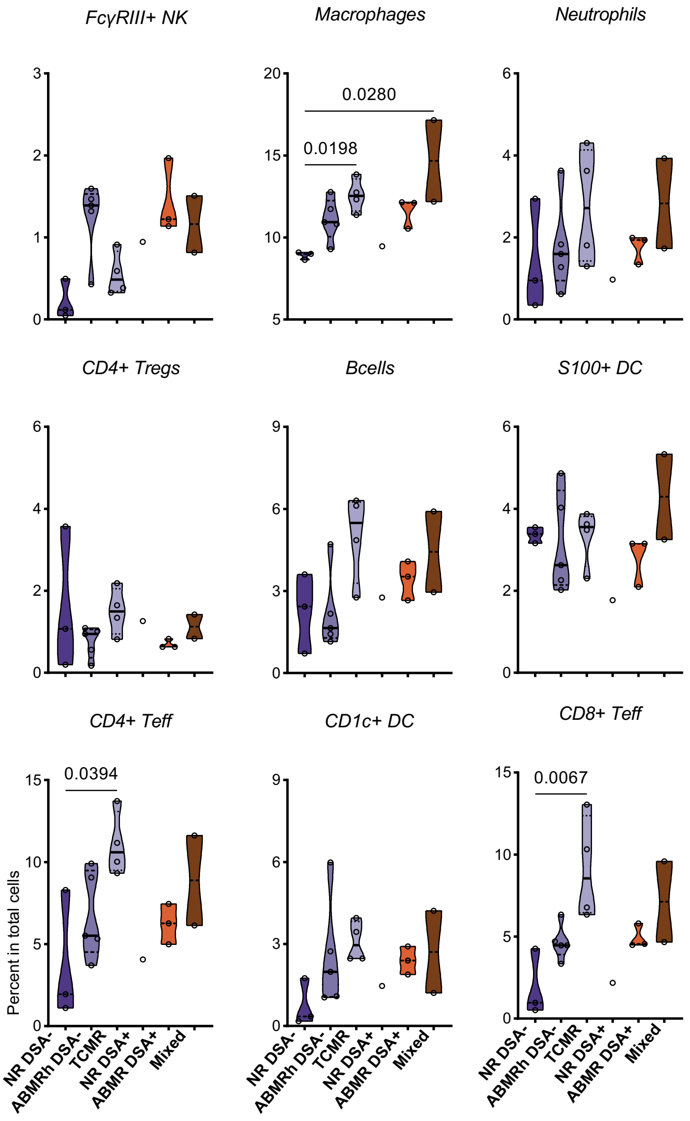

FcγRIII+ NK
Macrophages
Neutrophils
0.0280
0.0198
CD4+ Tregs
Bcells
S100+ DC
CD4+ Teff
CD1c+ DC
CD8+ Teff
0.0394
0.0067
Percent in total cells
NR DSA-
ABMRh DSA-
TCMR
NR DSA+
ABMR DSA+
Mixed

Supplementary Fig. 5

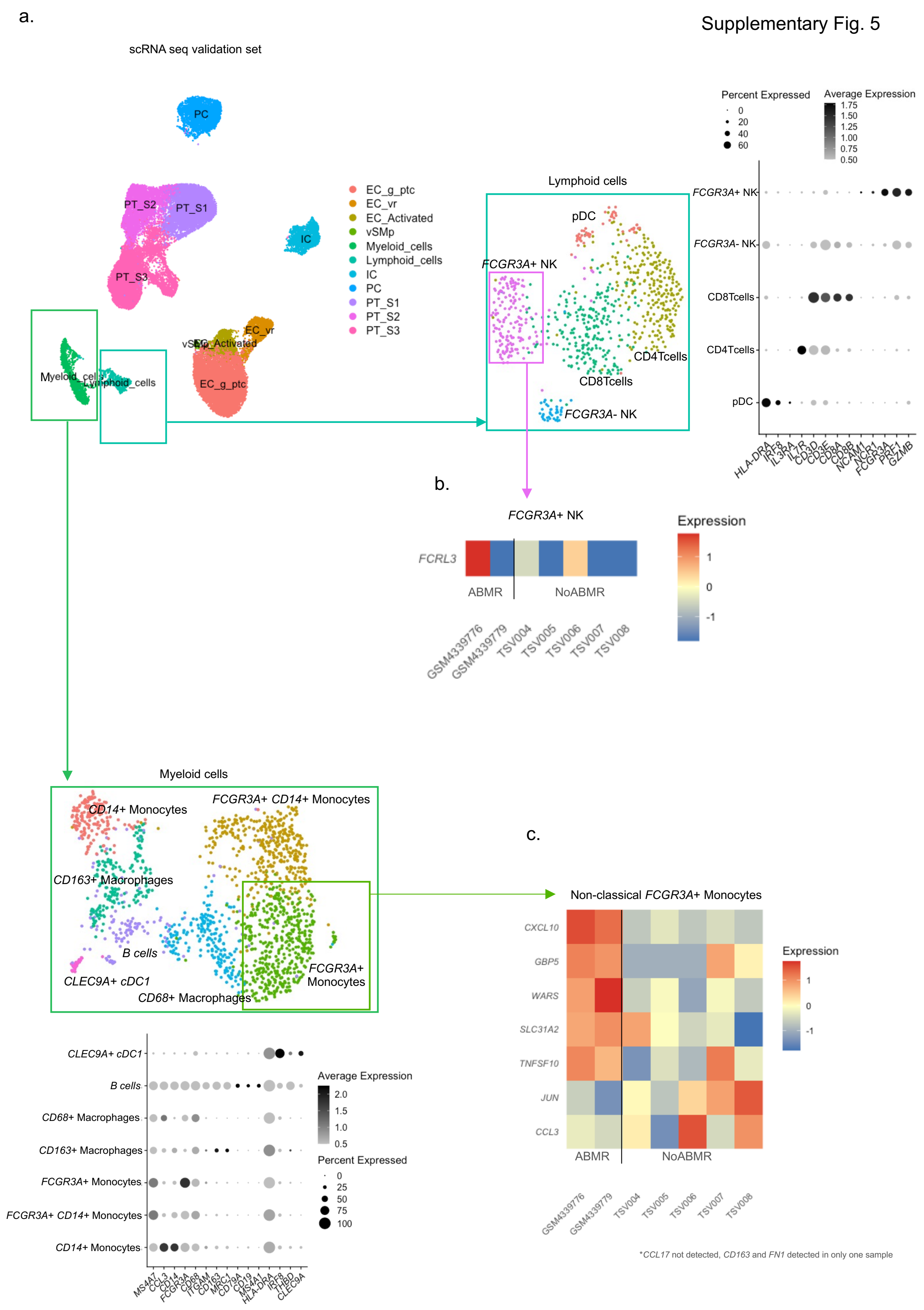

Supplementary Fig. 6

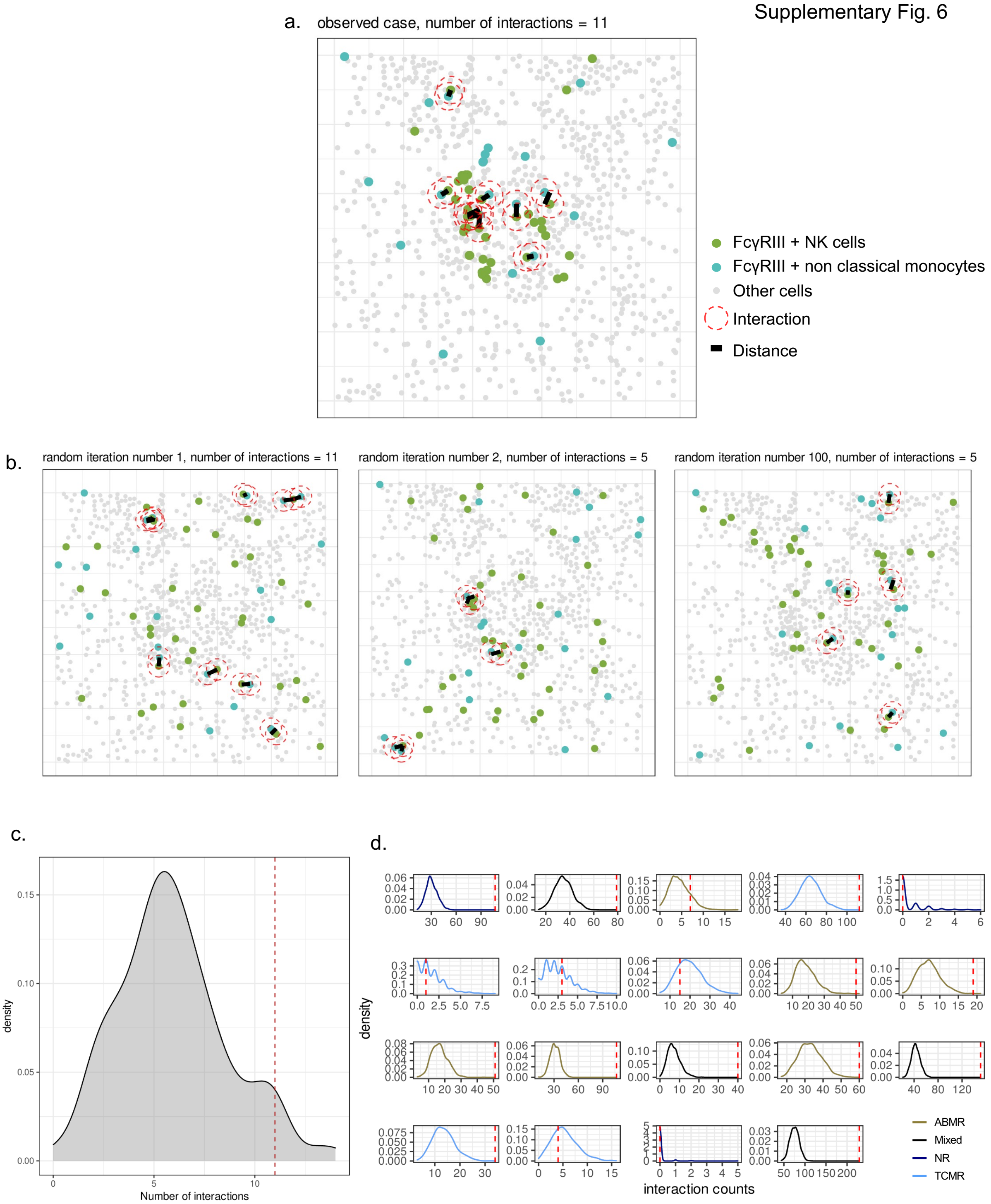

**Supplementary figure Legends**

Fig S1: a) Quality control and parameters of the generation of the single cell dataset b) UMAP visualization of the whole dataset grouped by original biopsy identity. c) UMAP visualization of each individual biopsy in the dataset demonstrating equal coverage of all cell clusters

Fig S2: a) UMAP visualization of the cells corresponding to the five biopsies derived from a female donor and a male recipient b) Dot plot showing the expression of sex-related genes. a) UMAP visualization of the cells corresponding to the single biopsy derived from a male donor and a female recipient b) Dot plot showing the expression of sex-related genes.

Fig S3: a) Correlation of deconvoluted cell populations based on pseudobulk analysis of single cell RNA seq samples (n=16) and actual cell counts in the sample. b) Correlation as described in a) for broader defined cell types. c) Workflow of OPAL immunofluorescence imaging and subsequent computerized segmentation, phenotyping and quantification. d) Correlation between the deconvoluted cell populations based on microarray data and the quantified populations using OPAL imaging.

Fig S4: Violin plots depicting the proportion of indicated cells measured by MILAN method regarding clinical outcome.

Fig S5: Single cell validation set. a) Public data derived from kidney biopsies were analyzed as previously described [3–5]. Lymphoid and myeloid cells were subclustered and reintegrated. (b) Heatmap showing the expression of indicated gene in *FCGR3A*+ NK is depicted. (c) Heatmap showing the expression of indicated genes in *FCGR3A*+ *CD14*- monocytes is depicted.

Fig S6: Neighborhood analysis: intermediate results. a) Scatter plot showing the location of FcγRIII + NK cells (green), FcγRIII + non classical monocytes (turquoise), and other cell types (gray) in a preselected tissue area for the observed data. FcγRIII + NK cells in the neighborhood of FcγRIII + non classical monocytes are represented by red dashed circles and connecting black lines. b) The same tissue area is represented after randomly permutating the label of each celltype. The reader should note that both the location of each cell as well as the number of cells of each type have been preserved. Only their labels have been randomly permutated. This process is repeated 1000 times. Here iteration numbers 1 (left), 2 (center), and 100 (right) are represented. c) Density plot showing the distribution of counts found in all 1000 random iterations. The vertical red dashed line instead represents the number of counts found in the observed data. We relative number of random cases with more counts than the observed cases can be interpreted as an empirical p-value representing the significance of a cell-cell interaction. d) The same exercise was repeated for each individual sample taking into account the whole tissue. These density plots represent an intermediate step towards the plot shown in Figure 9C.
